# Implementation of an Intervention to Improve Blood Culture Stewardship in 14 Adult ICUs and Wards

**DOI:** 10.64898/2026.10.29.26366841

**Authors:** Valeria Fabre, Yea-Jen Hsu, Anping Xie, Alejandra B. Salinas, Pilar Beccar-Varela, Viviana Martinez, Kathleen O. Degnan, Erin B. Gettler, Laurel J. Glaser, J. Kristie Johnson, Rebekah W. Moehring, Guillermo Rodriguez-Nava, Jonathan H. Ryder, Adam Ressler, Jorge L. Salinas, Gregory M. Schrank, Thomas R. Talbot, Trevor C. Van Schooneveld, Anastasia Wasylyshyn, Sara E. Cosgrove

## Abstract

**Importance:** Data are limited on factors associated with successful implementation of diagnostic stewardship interventions to improve blood culture (BCx) ordering in adult inpatients.

**Objective:** To evaluate the impact of a quality improvement (QI) initiative to improve BCx ordering and to evaluate factors associated with improved practices.

**Design, Setting, and Participants:** We conducted a pragmatic QI initiative over a one-year period in seven acute care academic hospitals in the United States. A total of 14 adult units, seven intensive care units (ICUs) and seven wards from each hospital were included. The study was conducted between April ^st^ 2022 and June 30^th^, 2024. Data analysis was performed March 1^st^ to October 25^th^, 2026.

**Intervention:** The QI included implementation of an evidence-based BCx algorithm with indications for blood cultures, education on the algorithm, and monitoring and feedback of BCx practices.

**Outcomes and Measures:** The primary outcome was BCx utilization (BCU) 12 months before and after implementation. Using the Consolidated Framework for Implementation Research, we assessed implementation through interviews with QI teams, diaries of QI activities completed by each team, and a survey of frontline providers. Coincidence analysis was used to identify implementation factors associated with improvement in BCU.

**Results:** Four ICUs had significantly lower BCU (∼20% reduction), one of them also had a 50% increase in BCx positivity. Greater baseline BCx inappropriateness was associated with significant reduction in BCU. Two wards had a significant increase in BCU, one of which had a concomitant increase in BCx positivity. An additional ward had a 60% increase in BCx positivity despite similar BCU before and after QI implementation. Coincidence analysis identified several factors associated with improved BCU, including perceptions that the algorithm was easy to use and reduced unnecessary repeat BCx, as well as nurse inclusion in educational activities and regular education and feedback on BCx practices.

**Conclusion:** Baseline BCU should inform the design and focus of BCx stewardship initiatives. Effective improvement efforts require multifaceted implementation strategies.

**Key Points:** *Question:* Is a blood culture stewardship initiative associated with improved blood culture use in intensive care units (ICUs) and wards, and what drives success?

*Findings:* In this multicenter study of 14 ICUs and wards, an algorithm with education and feedback was associated with ∼20% lower blood culture use in 4 ICUs and up to 60% higher positivity in 1 ICU and 2 wards. Success was linked to higher baseline inappropriate use, nursing education, and provider perception that the algorithm was helpful and easy to use.

*Meaning:* The intervention works best in units with higher inappropriate use, supported by nursing engagement and clinician-centered education.

## Introduction

Previous studies from the United States and elsewhere have shown that as many as 60% of blood cultures (BCx) may be inappropriate on the basis of indication^1–3^. BCx stewardship initiatives aimed at improving ordering practices have been associated with increased BCx positivity (i.e., bacteremia detection), reduced antibiotic use, and fewer central-line associated bloodstream infections (CLABSIs)^1,4^.

We previously developed and implemented an evidence-based clinical decision support algorithm to help clinicians identify conditions associated with high probability of bacteremia as well as those in which BCx have low yield in the intensive care unit (ICU) and general ward settings^1,5^. This approach has since been adapted and implemented in other settings including the Emergency Department and across health systems, with variable results, suggesting that implementation characteristics may affect outcomes^6,7^. Moreover, most of these studies were limited to single hospitals or single health systems and did not examine implementation characteristics. To address these gaps, we conducted a BCx stewardship quality improvement (QI) initiative focused on indications for BCx use and evaluated the implementation to characterize factors associated with desired outcomes across 14 units in seven U.S. academic hospitals.

## Methods

### Study participants and study design

We conducted a multicenter QI project focused on optimizing BCx indications among patients admitted to adult ICUs and wards across seven academic hospitals in the U.S. This study was conducted as part of a Centers for Disease Prevention and Control Prevention Epicenter Program (CDC-PEP)-funded collaborative to evaluate blood culture utilization (BCU) rates in U.S. medical ICUs and wards^8^. Hospitals were recruited through open calls via the Society of Healthcare Research Network (SHEA-RN) and through CDC-PEP calls. Small hospitals with ≤50 beds were excluded. Basic characteristics of participating hospitals are shown in **eTable** 1 in the Supplement.

The study had two aims: first, to evaluate the impact of the QI initiative on BCU; and second, to assess its implementation. For aim 1, we used a 12-month pre-post study design with the primary outcome being the number of BCx collected per 1,000 patient-days (PD). Additional outcomes are described below). For Aim 2, we used a multimodal approach informed by the Consolidated Framework for Implementation Research (CFIR)^9^, including interviews with QI teams, survey to front-line providers, and diaries of activity regularly maintained by each hospital’s QI team. Additional details regarding the QI initiative and analytical methods are provided below. This study was approved by the Johns Hopkins Medicine Institutional Review Board as Not Human Subjects Research (NHSR)/Quality Improvement (QI).

### QI implementation approach

The BCx stewardship QI included three main components: a) adaptation and implementation of a published evidence-based BCx algorithm^5^; b) bedside provider education on the contents of the BCx algorithm by local QI teams; and c) feedback on BCU and/or BCx inappropriateness by local QI teams. Each hospital developed a QI team to lead implementation of the QI. The QI team was composed by experts in hospital epidemiology/infection prevention, antimicrobial stewardship, or microbiology. Each QI team was responsible for selecting one intensive care unit (ICU) and one ward to participate, implementing the QI intervention, and collecting and submitting data to the coordinating center (JHU).

Each team adapted the BCx algorithm in consultation with participating units’ thought leaders (adaptations to the algorithm by each site can be found in **eTable 2** in the Supplement). Additionally, each QI team retrospectively reviewed and adjudicated a random sample of BCx to be able to provide feedback on BCx use to frontline providers using a standardized tool in REDCap. The exact number of cases adjudicated, and timeline are shown in **eTables 3 and 4** in the Supplement. The frequency and mode of educational or feedback delivery were determined by each local QI team according to local workflows. Individual dates for implementation of the QI at each site are shown in **eTable 1** in the Supplement. The JHU team supported participating sites by providing implementation resources, including educational materials on BCx, an editable version of the BCx algorithm, and a data-tracking tool to monitor BCU rates. In addition, the JHU team provided ongoing coaching through monthly check-in calls throughout the study period and conducted the data analyses. There were no BCx supply shortages experienced by sites during the study period.

### Evaluation of the impact of the QI on BCU

The primary outcome was BCx utilization (BCU) which was measured as the number of BCx collected/1,000 patient-days (PD) during the 12 months before and after the implementation of the QI. Process measures included single BCx (i.e., number of BCx collected as a single set in a 24 h period), BCx positivity (measured as the number of positive BCx excluding contaminants divided by all BCx collected), and BC contamination (BCC) rates as previously defined^10^. BCx intensity was measured as the number of BCx divided by number of admissions with at least one BCx^11^.

Clinical outcomes included length of stay, antibiotic days of therapy (DOT), and CLABSI rates as defined by NHSN. Safety measures included: hospital compliance with the Severe Sepsis and Septic Shock Management bundle (SEP-1), and all-cause in-unit mortality. Data were extracted electronically at each site from the relevant source, depending on the outcome (e.g., EHR, NHSN, REDCap). We estimated incidence rate ratios (IRR) with 95% confidence intervals (CI) to evaluate differences in BCU before and after the QI using Poisson regression. The association between baseline BCx inappropriateness and BCU was measured using negative binomial regression model, treating inappropriateness as a continuous variable. Antibiotic use and CLABSI rates were analyzed similarly using Poisson regression. We calculated odds ratio (OR) with 95% CI using logistic regression models to evaluate BCx positivity, BCC, single BCx, and BCx intensity before and after the QI. Each hospital was assigned a letter for reporting purposes, and the ICU and ward labeled with the same letter represent the same institution. We used STATA v18.0 (StataCorp, College Station, TX).

### Evaluation of the QI implementation

Guided by the CFIR, we conducted semi-structured interviews with QI teams to explore perceived barriers and facilitators to BCx stewardship. Each team also completed a structured quarterly activity diary housed in REDCap, and bedside providers completed an electronic survey on their perceptions regarding the BCx algorithm, BCx practices, and the QI. Interviews and surveys took place 6–9 months after implementation (the interview guide and questionnaire are provided in **eMethods** and **eTable 5** in the Supplement). Interview participants provided oral consent, and survey completion was taken as consent. Two investigators (AX, VF) independently reviewed each transcript for recurring ideas and concepts and reached consensus on shared themes. Survey and diary data were summarized descriptively.

To identify implementation factors associated with lower BCU, we performed a coincidence analysis, an approach previously used to evaluate QI implementation^12^. This configurational comparative method, grounded in set theory and Boolean logic, accommodates multiple factors and identifies minimally sufficient and necessary conditions (MSCs) from observed configurations^13^. Three investigators (VF, YJH, AX) with expertise in infectious diseases, human factors engineering, health services research, and statistics selected candidate factors based on expert consensus from diaries of activities and the healthcare worker survey. The combination of factors that explain the outcome is called a “solution”. We report the coverage for each MSC defined as the percent of hospitals with the outcome explained by the factor. We conducted a sensitivity analysis accounting for an increase in BCx positivity without a reduction in BCU, given that no significant reduction in BCU was observed in the wards. Analyses were conducted in R using the cna package (version 4.0.3).

### Reporting

We followed the Standards for Quality Improvement Reporting Excellence (SQUIRE) 2.0 guidelines^14^ for reporting (**eAppendix in the Supplement**).

## Results

### Characteristics of participating units and baseline BCx parameters

Seven ICUs (five medical, one surgical and one cardiac) and seven wards (five medical, one medical-surgical and one cardiac) from unique hospitals were included in the analysis. Their baseline BCx parameters (BCU, BCx positivity, BCC and BCx inappropriateness) are summarized in **Table 1**. The mean BCU for the ICUs ranged from 128 to 380 BCx/1,000 PD, and for the wards, from 33 to 84 BCx/1,000 PD. Mean true BCx positivity across sites ranged from 4.9% to 21.5% for ICUs, and from 0% to 20% for the wards. Overall, the proportion of inappropriate BCx at the time of the QI implementation was 27% for ICUs (range, 15% - 50%) and 39% for the wards (range, 20% - 67%). Top reasons for inappropriate BCx included persistent leukocytosis or fever and negative BCx within the last 72 hours, isolated fever or leukocytosis, and non-infectious presentations (**Table 2** and **eTables 3 and 4 in the Supplement**).

**Table 1.** Baseline blood culture (BCx) quality indicators for intensive care units (ICU) and wards at participating sites.

| **Unit** | **Unit type\*** | **Mean BCx/1,000 patient days (95% CI)** | **Mean BCx positivity (%)** | **Mean single BCx (%)** | **Mean BCC (%)** | **Inappropriate BCx (%)** |
|---|---|---|---|---|---|---|
| ICU A | CC:M | 248 (238 – 257) | 10.2 | 8.9 | 1.4 | 16 |
| ICU B | CC:M | 192 (183 – 201) | 4.9 | 9.2 | 2.4 | 30 |
| ICU C | CC:M | 380 (364 – 395) | 8.4 | 5.5 | 1.4 | 33 |
| ICU D | CC:M | 316 (297 – 336) | 8.7 | 1.6 | 2.1 | 50 |
| ICU E | CC:S | 238 (226 – 250) | 21.5 | 13.9 | 3.5 | 15 |
| ICU F | CC:M | 201 (191 – 213) | 6.4 | 0.8 | 0.5 | 37 |
| ICU G | CC:C | 128 (122 – 133) | 9.6 | 4.1 | 2.7 | 20 |
| Ward A | Ward:M | 71 (65 – 76) | 9.3 | 5.8 | 1.4 | 20 |
| Ward B | Ward:M | 33 (27 – 40) | 0 | 9.2 | 0 | 50 |
| Ward C | Ward:M | 84 (79 – 90) | 9.3 | 5.5 | 0.9 | 30 |
| Ward D | Ward:MS | 58 (54 – 62) | 7.6 | 1.6 | 2.2 | 40 |
| Ward E | Ward: M | 82 (79 – 86) | 20.0 | 6.5 | 0.3 | 50 |
| Ward F | Ward:M | 68 (65 – 72) | 10.3 | 1.1 | 0.9 | 67 |
| Ward G | Ward:C | 51 (47 – 56) | 6.6 | 5.1 | 2.8 | 60 |

\*Based on the National Healthcare Safety Network (NHSN) designation (CC:M, medical ICU; CC:C cardiac ICU; Ward: M, medicine ward, Ward:MS, medical-surgical ward; Ward:C, cardiac ward), BCC: blood culture contamination.

**Table 2.**
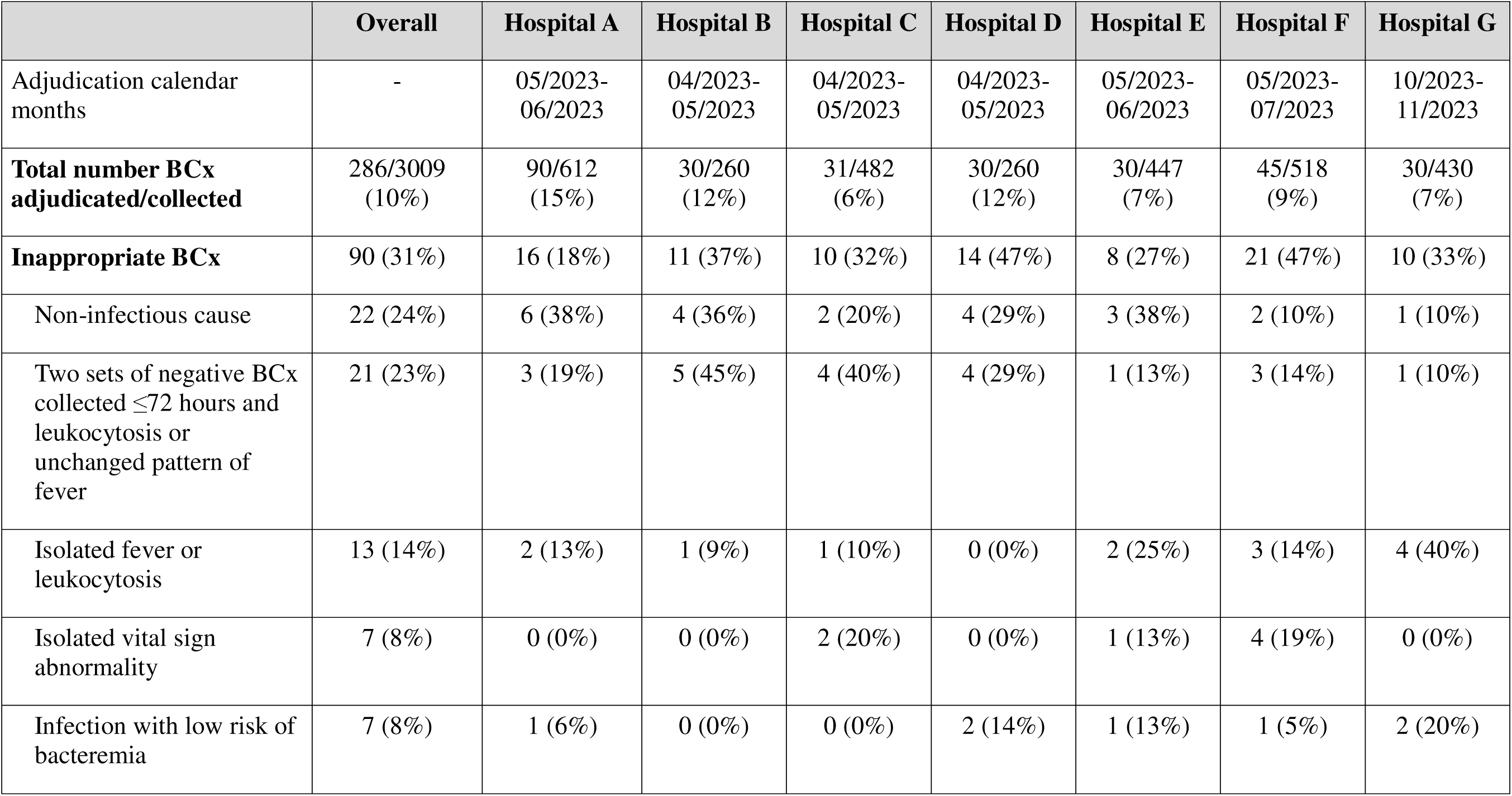
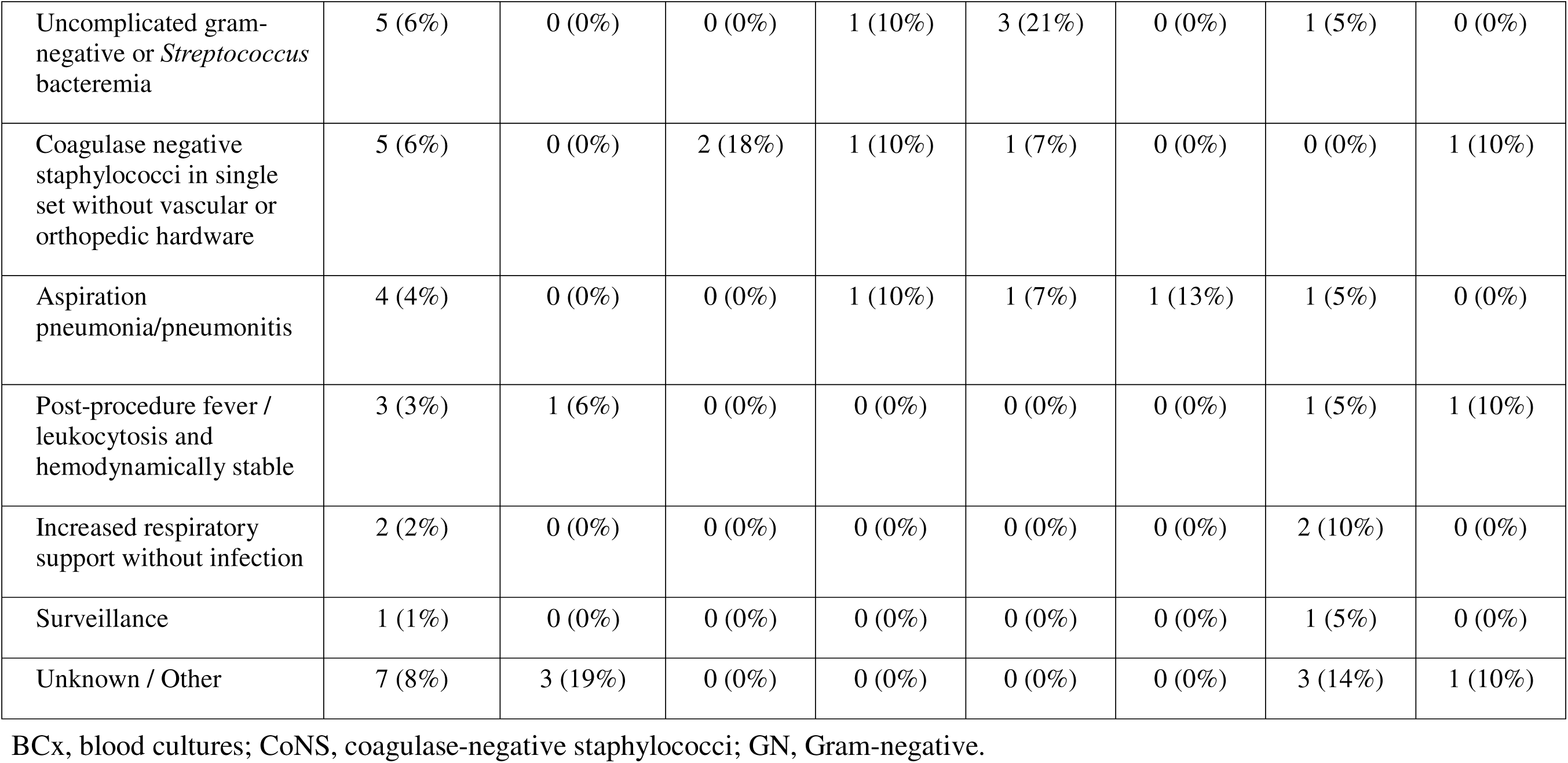
Inappropriate blood cultures (BCx) indications by study site. Blood cultures collected in participating units during the first two months of the BCx Stewardship Quality Improvement (QI) initiative were adjudicated retrospectively by each local QI team through chart review, using the adapted BCx algorithm as the reference standard. A BCx may have more than one reason, so reason percentages may not sum to 100%.

### Impact of the BCx Stewardship QI in ICUs

The impact of the BCx stewardship QI on BCU is presented in **Table 3**. BCU decreased by ∼20% in four ICUs (ICUs B, C, D, and F) and remained unchanged in three others (ICU A, E, and G). Higher BCx inappropriateness at baseline was associated with a significant reduction in BCU (IRR 0.81, 95% CI 0.71 – 0.93, *P*=0.002). BCx positivity remained similar in all ICUs except ICU B (50% higher) and E (50% lower). BCx intensity was lower in ICUs C and D (**eTable 6 in the Supplement**). Single BCx rates were higher post QI implementation in ICU D. The proportion of BCC remained similar between periods across ICUs (**eTable 7 in the Supplement**). Clinical outcomes before and after the QI were largely unchanged except for antibiotic use. IV Vancomycin use was lower in the post-implementation period for all units, while use of broad-spectrum β-lactams remains unchanged in most units, lower in ICU A and ICU G, and higher in ICU D (**eTables 8-10 in the Supplement**).

**Table 3.**
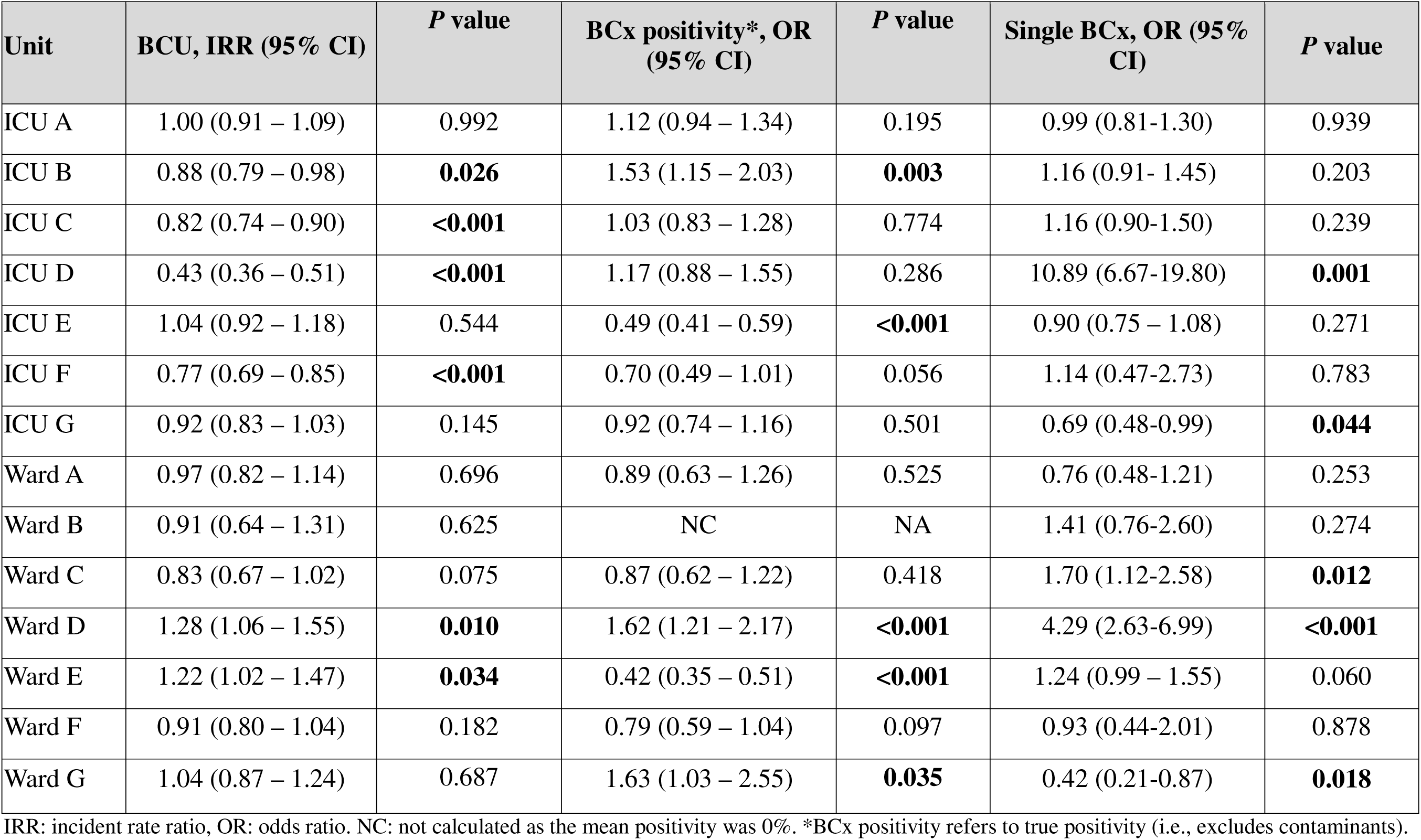
Impact of the blood culture stewardship quality improvement (QI) initiative on blood culture (BCx) quality indicators. Blood culture utilization (BCU), BCx positivity, and single BCx rates were compared 12 months before and after the QI implementation.

| **Unit** | **BCU, IRR (95% CI)** | ***P* value** | **BCx positivity\*, OR (95% CI)** | ***P* value** | **Single BCx, OR (95% CI)** | ***P* value** |
|---|---|---|---|---|---|---|
| ICU A | 1.00 (0.91 – 1.09) | 0.992 | 1.12 (0.94 – 1.34) | 0.195 | 0.99 (0.81-1.30) | 0.939 |
| ICU B | 0.88 (0.79 – 0.98) | **0.026** | 1.53 (1.15 – 2.03) | **0.003** | 1.16 (0.91- 1.45) | 0.203 |
| ICU C | 0.82 (0.74 – 0.90) | **<0.001** | 1.03 (0.83 – 1.28) | 0.774 | 1.16 (0.90-1.50) | 0.239 |
| ICU D | 0.43 (0.36 – 0.51) | **<0.001** | 1.17 (0.88 – 1.55) | 0.286 | 10.89 (6.67-19.80) | **0.001** |
| ICU E | 1.04 (0.92 – 1.18) | 0.544 | 0.49 (0.41 – 0.59) | **<0.001** | 0.90 (0.75 – 1.08) | 0.271 |
| ICU F | 0.77 (0.69 – 0.85) | **<0.001** | 0.70 (0.49 – 1.01) | 0.056 | 1.14 (0.47-2.73) | 0.783 |
| ICU G | 0.92 (0.83 – 1.03) | 0.145 | 0.92 (0.74 – 1.16) | 0.501 | 0.69 (0.48-0.99) | **0.044** |
| Ward A | 0.97 (0.82 – 1.14) | 0.696 | 0.89 (0.63 – 1.26) | 0.525 | 0.76 (0.48-1.21) | 0.253 |
| Ward B | 0.91 (0.64 – 1.31) | 0.625 | NC | NA | 1.41 (0.76-2.60) | 0.274 |
| Ward C | 0.83 (0.67 – 1.02) | 0.075 | 0.87 (0.62 – 1.22) | 0.418 | 1.70 (1.12-2.58) | **0.012** |
| Ward D | 1.28 (1.06 – 1.55) | **0.010** | 1.62 (1.21 – 2.17) | **<0.001** | 4.29 (2.63-6.99) | **<0.001** |
| Ward E | 1.22 (1.02 – 1.47) | **0.034** | 0.42 (0.35 – 0.51) | **<0.001** | 1.24 (0.99 – 1.55) | 0.060 |
| Ward F | 0.91 (0.80 – 1.04) | 0.182 | 0.79 (0.59 – 1.04) | 0.097 | 0.93 (0.44-2.01) | 0.878 |
| Ward G | 1.04 (0.87 – 1.24) | 0.687 | 1.63 (1.03 – 2.55) | **0.035** | 0.42 (0.21-0.87) | **0.018** |

IRR: incident rate ratio, OR: odds ratio. NC: not calculated as the mean positivity was 0%. \*BCx positivity refers to true positivity (i.e., excludes contaminants).

### Impact of the BCx Stewardship in wards

No ward showed a significant decrease in BCU. Wards D and E demonstrated significant increases in BCU, accompanied by an increase in BCx positivity in Ward D and a decrease in BCx positivity in Ward E. In Ward G, BCx positivity increased after QI implementation, along with a lower rate of single BCx collections. BCx intensity was lower in wards C, D, and F (**eTable 6 in the Supplement**). BCC rates as well as clinical outcomes were largely similar before and after the QI (**eTables 7-10 in the Supplement**).

### QI implementation evaluation

A summary of implementation activities is shown in **eTable 11** in the Supplement. QI teams in hospitals A and D reported the highest number of implementations. There were differences between sites regarding feedback and sustained education (e.g., Hospital E gave no feedback on BCU or appropriateness after implementation, and nurses were included in education everywhere except hospitals A and E).

Twenty-six individuals involved in the QI, including participating unit medical directors, unit champions, infection preventionists and microbiologists, participated in semi-structured interviews. They identified the following facilitators to the QI implementation: prior experience with diagnostic stewardship, a supportive and engaged physician unit director, and nursing engagement. Lower use of BCx in the wards compared to the ICU, the need to train users on how to apply the algorithm, the inability to efficiently assess compliance with the BCx algorithm, and conflicts around pressure to expedite decisions about BCx by sepsis initiatives were perceived as barriers to the QI. Additional information from interviews is presented in **eTable 12** in the Supplement.

Two hundred and twelve frontline providers participated in a survey regarding the BCx stewardship QI (detailed responses in **eTables 5 and 13** in the Supplement). Most (68%) participants were familiar with the QI. Among these, general perceptions about the BCx algorithm were positive. Overall, most individuals perceived the BCx algorithm to be applicable to most patients in their units (86% and 89% of respondents agreed with this for the ICU and wards, respectively). More participants agreed that the BCx algorithm was more useful to reduce unnecessary repeat BCx than to reduce BCx for isolated fever or leukocytosis (73% and 64%, respectively). There was an overall perception that the algorithm is easy to use, although with variability across sites (e.g., 57% of participants in hospital A versus >90% of participants in hospitals B and D found it easy to use).

In the coincidence analysis, nine solutions were identified (**Table 4, eTables 14-22** in the Supplement). All solutions demonstrated 100% consistency, indicating that the improved outcome occurred whenever the solution was present, and 100% coverage, indicating that all ICUs with improved outcomes were explained by the solution. With the exception of Solution 1, which includes two factors, each solution included three factors. Solutions 2 through 7 each included Factor 3 (frontline workers reporting higher perceived helpfulness of the BCx algorithm in reducing unnecessary repeat BCx) combined with another factor related to continuous nursing education, greater awareness of the QI initiative, or higher perceived ease of use of the BCx algorithm. These were among the strongest MSCs and explained three of the four hospitals with improved BCU (coverage = 75%). Results remained similar on sensitivity analysis of increased positivity as improved outcome.

**Table 4.** Factors associated with lower Blood Culture Utilization (BCU) in Intensive care Units based Coincidence Analysis. Data source included diary of activities (D) and surveys to frontline providers (S). Each “Solution” represents a distinct combination of factors associated with lower BCU and should be read vertically as a configuration.

|  | **Solution** |  |  |  |  |  |  |  |  |
|---|---|---|---|---|---|---|---|---|---|
| **Factor in Solution** | **1** | **2** | **3** | **4** | **5** | **6** | **7** | **8** | **9** |
| Feedback to providers on BCU before QI implementation (D) | ⊕ |  |  |  | + | + | + |  |  |
| Reported resistance to use the BCx algorithm (S) | ⊞ |  |  |  |  |  |  | + | + |
| Perceptions that the algorithm helps reduce unnecessary BCx (S) |  | ⊕ | ⊕ | ⊕ | ⊕ | ⊕ | ⊕ |  |  |
| Nurses included in educational activities (D) |  | ⊕ |  |  | ⊕ |  |  | ⊕ |  |
| Providers’ awareness of the BCx Stewardship QI (S) |  |  | ⊕ |  |  | ⊕ |  |  |  |
| Perceptions that the algorithm is easy to use in practice (S) |  |  |  | ⊕ |  |  | ⊕ |  |  |
| Education to providers on BCU before QI implementation (D) |  | – | – | – |  |  |  |  |  |
| Regular education on BCx practices after QI implementation (D) |  |  |  |  |  |  |  | ⊕ | ⊕ |
| Regular feedback on BCU during QI (D) |  |  |  |  |  |  |  |  | ⊕ |
| Consistency of solution (percent of hospitals with the solution that improved BCU) | 100% | 100% | 100% | 100% | 100% | 100% | 100% | 100% | 100% |
| Coverage of solution (percent of hospitals with improved BCU explained by the MSC or solution) | 100% | 100% | 100% | 100% | 100% | 100% | 100% | 100% | 100% |
| Coverage of best-performing MSC in solution (percent of hospitals with improved BCU explained by the best performing component in the solution) | 50% | 75% | 75% | 75% | 75% | 75% | 75% | 75% | 75% |

Quality improvement (QI). ⊕ indicates that a factor is present and serves as a component of the stronger minimal sufficient condition (MSC) or “building block” of the solution; ⊞indicates that a factor is present and serves as another component of the solution; + indicates that a factor is present and serves as a component of the weaker MSC of the solution; – indicates that a factor is absent and serves as a component of the weaker MSC of the solution.

## Discussion

We implemented and evaluated a BCx diagnostic stewardship intervention across seven ICUs and seven wards. All units adopted a BCx algorithm to guide ordering decisions and delivered education and variable feedback on BCx use. The initiative was associated with some reduction in BCU among most units, and with increased BCx positivity in some. Lowered BCx rates were associated with a higher baseline rate of inappropriate BCx, the inclusion of nurses in educational activities, and frontline providers’ perception that the algorithm was easy to use and helpful in avoiding unnecessary repeat cultures.

BCx stewardship offers several benefits including reducing unnecessary antibiotic use and testing, lower BCC by limiting unwarranted BCx, and fewer misattributed CLABSIs^15^. Decision-support algorithms have shown to reduce unnecessary BCx and increasee BCx positivity without harming patients^1,16,17^, yet their impact varies widely across settings, and the determinants of successful implementation remain poorly understood. Ramos and colleagues observed differences in BCU across three EDs in one health system after adoption of a shared BCx algorithm^6^. The authors concluded this difference could be due to the intensity of feedback delivered by the QI team to frontline providers on BCx use (only weekly feedback was effective compared to bi-monthly or no feedback). Our study offers new information in identifying the factors that contribute to successful BCx stewardship implementation. We likewise found feedback to be important, although less intensive feedback sufficed. Interviews provided additional context for these findings. Feedback built around real cases, in which ordering providers critically appraise their own decisions, may be more effective than reporting BCx counts or offering passive education about bacteremia risk. These in-person case discussions increased trainees’ confidence in using the algorithm. Others have shown that while introducing BCx guidance on indications in the EMR reduced BCU during a bottle shortage, BCU rebounded once supplies recovered, underscoring the need for behavioral interventions to sustain change^18,19^.

Frontline awareness of the initiative, nursing education, and the algorithm’s perceived ease of use and helpfulness in reducing repeat cultures were also associated with improvement in our study. Nursing involvement mattered most in ICUs, where nurses collect BCx, know the patient’s clinical status and recent results in detail, and are strongly committed to CLABSI prevention. The intervention appeared less effective on wards, where BCx are ordered less often, phlebotomy services draw BCx, and nurses are less likely to question whether a culture is needed. Our finding that greater perceived helpfulness of the BCx algorithm in reducing unnecessary repeat cultures was associated with improved practice is not surprising, as these decisions are less ambiguous. Fever, by contrast, is a nonspecific sign, and patients without a clear source of infection may not fit neatly within the algorithm. In prior work, most clinicians did not consider BCx indicated for new-onset fever yet acknowledged that cultures are still routinely obtained in this setting^20,21^.

Benchmarks can help units identify under- or overutilization though remain investigational and preliminary. Among the four study units with a published reference benchmark for their unit type that did not show a reduction in BCU after the QI, all had BCU rates below that benchmark at baseline^8^. Ward D, for example, began below the previously reported benchmark of 65 BCx/1,000 patient-days for medical-surgical wards^8^; after the QI, its BCU rose by 28% and its positivity by 60%, suggesting prior underutilization. Furthermore, BCx intensity fell, suggesting the intervention reduced repeat BCx.

This study has limitations. All seven hospitals were academic medical centers, albeit in different regions. Sites were recruited for their interest and capacity to join a multicenter QI project with a tight timeline and may therefore be better resourced than other institutions. This may may limit generalizability of findings. We were unable to measure algorithm compliance, although we assessed adoption and usability (e.g., applicability to the unit population, ease of use). Coincidence analysis offers a robust structured approach to identifying implementation factors; however, our small sample may have obscured other influences on outcomes. We focused on the impact of the QI on 12 months post implementation. Future studies examining sustainability of BCx stewardship initiatives are needed. Appropriateness review was performed on a subset of cases because this process is resource-intensive. Future work should focus on developing tools to support efficient adjudication of BCx^22^. We did not collect patient level data and therefore could not account for any changes in case mix between study periods. Finally, because BCC is expressed as a percentage of BCx collected, our results may understate the QI’s effect on contamination.

In summary, a BCx algorithm can be a useful tool for optimizing BCx use, but meaningful change requires training to help providers apply the algorithm to individual patients, as well as nursing engagement to prompt discussion about the necessity of BCx.

## Acknowledgements

The authors would like to thank the following individuals for their contributions to the implementation of the QI, Carlo Gentleman, Siu Yan A. Yeung, PharmD, Tania Rodriguez, Leigh Cressman, Edward Chen, and Maiko Suarez.

## Author Contributions

Concept and design: Fabre, Cosgrove.

Acquisition, analysis, or interpretation of data: All authors.

Critical revision of the manuscript for important intellectual content: All authors.

Statistical analysis: Hsu, Xie, Fabre.

Obtained funding: Fabre, Cosgrove.

Administrative, technical, or material support: Salinas, Martinex, Beccar-Varela.

## Disclaimer

The content is solely the responsibility of the authors and does not necessarily represent the official view of the CDC.

## Data availability statement

All data produced in the present work are contained in the manuscript.

## References

1. Fabre V, Klein E, Salinas AB, et al. A Diagnostic Stewardship Intervention To Improve Blood Culture Use among Adult Nonneutropenic Inpatients: the DISTRIBUTE Study. J Clin Microbiol. 2020;58(10).

2. Siev A, Levy E, Chen JT, et al. Assessing a standardized decision-making algorithm for blood culture collection in the intensive care unit. J Crit Care. 2023;75:154255.

3. Drager S, Giehl C, Sogaard KK, et al. Do we need blood culture stewardship programs? A quality control study and survey to assess the appropriateness of blood culture collection and the knowledge and attitudes among physicians in Swiss hospitals. Eur J Intern Med. 2022;103:50–56.

4. Woods-Hill CZ, Colantuoni EA, Koontz DW, et al. Association of Diagnostic Stewardship for Blood Cultures in Critically Ill Children With Culture Rates, Antibiotic Use, and Patient Outcomes: Results of the Bright STAR Collaborative. JAMA Pediatr. 2022;176(7):690–698.

5. Fabre V, Sharara SL, Salinas AB, Carroll KC, Desai S, Cosgrove SE. Does This Patient Need Blood Cultures? A Scoping Review of Indications for Blood Cultures in Adult Nonneutropenic Inpatients. Clin Infect Dis. 2020;71(5):1339–1347.

6. Ramos J, Theophanous R, Gettler E, et al. A comparison of blood culture diagnostic stewardship across three emergency departments in a healthcare network. Am J Emerg Med. 2025;93:135–139.

7. Takkavatakarn K, Patel G, Oh W, et al. Electronic clinical decision support system guided blood culture stewardship in emergency departments: response to the national blood culture media shortage. Infect Control Hosp Epidemiol. 2025;46(6):650–653.

8. Fabre V, Hsu YJ, Carroll KC, et al. Blood Culture Use in Medical and Surgical Intensive Care Units and Wards. JAMA Netw Open. 2025;8(1):e2454738.

9. Damschroder LJ, Aron DC, Keith RE, Kirsh SR, Alexander JA, Lowery JC. Fostering implementation of health services research findings into practice: a consolidated framework for advancing implementation science. Implement Sci. 2009;4:50.

10. Fabre V, Hsu YJ, Carroll KC, et al. Multicenter evaluation of blood culture contamination and blood cultures practices in US acute care hospitals: time for standardization. J Clin Microbiol. 2025:e0053025.

11. Yu KC, Ye G, Edwards JR, et al. Hospital-onset bacteremia and fungemia: An evaluation of predictors and feasibility of benchmarking comparing two risk-adjusted models among 267 hospitals. Infect Control Hosp Epidemiol. 2022;43(10):1317–1325.

12. Dy SM, Acton RM, Yuan CT, et al. Association of Implementation and Social Network Factors With Patient Safety Culture in Medical Homes: A Coincidence Analysis. J Patient Saf. 2022;18(1):e249–e256.

13. Baumgartner M AM. Causal modeling with multi-value and fuzzy-set Coincidence Analysis. Political Science Research and Methods. 2020;8:526–542.

14. Ogrinc G, Davies L, Goodman D, Batalden P, Davidoff F, Stevens D. SQUIRE 2.0 (Standards for QUality Improvement Reporting Excellence): revised publication guidelines from a detailed consensus process. BMJ Qual Saf. 2016;25(12):986–992.

15. Fabre V, Carroll KC, Cosgrove SE. Blood Culture Utilization in the Hospital Setting: a Call for Diagnostic Stewardship. J Clin Microbiol. 2022;60(3):e0100521.

16. Wang MC, Zhou KJ, Shay SL, et al. The impact of a blood-culture diagnostic stewardship intervention on utilization rates and antimicrobial stewardship. Infect Control Hosp Epidemiol. 2024;45(5):670–673.

17. Seidelman JL, Moehring R, Gettler E, et al. Implementation of a diagnostic stewardship intervention to improve blood-culture utilization in 2 surgical ICUs: Time for a blood-culture change. Infect Control Hosp Epidemiol. 2024;45(4):452–458.

18. Shelly MJ, Kunz A, Mittal J, Corwin DS, Chapman C, Ender PT. Efficacy of a Multicenter Hospital Network’s Approach to Enacting Blood Culture Stewardship During a Global Shortage. Open Forum Infect Dis. 2025;12(6):ofaf294.

19. Ezran C, Herrle E, Yen CF, Mercuro NJ, Diekema DJ, Gordon LB. Shortage as a catalyst for high-value care: Evaluation of a blood culture stewardship intervention driven by supply chain disruption. J Hosp Med. 2026;21(4):368–374.

20. Fabre V, Cosgrove SE, Milstone AM, et al. Mixed-methods multicenter assessment of healthcare workers’ knowledge, perceptions, and practices related to blood culture utilization in hospitalized adults. Infect Control Hosp Epidemiol. 2024:1–9.

21. Fabre V, Milstone AM, Keller SC, Carroll KC, Cosgrove SE. Prescribers’ knowledge, attitudes and perceptions about blood culturing practices for adult hospitalized patients: a call for action. Infect Control Hosp Epidemiol. 2018;39(11):1394–1396.

22. Rodriguez-Nava G, Keyes T, Ambers N, et al. Using secure artificial intelligence agents integrated within the electronic medical record for the evaluation of blood culture appropriateness-Northern California, 2025. Infect Control Hosp Epidemiol. 2026;47(2):206–209.

